# CHWs’ and Health Personnel Perspectives in Yellow Fever Surveillance: A Qualitative Study in Two Health Districts of the Northwest Region of Cameroon

**DOI:** 10.1101/2025.08.28.25334621

**Authors:** Ngem Bede Yong, Nguemaïm Ngoufo Flore, Loveline Lum Niba, Gerald Ngo Teke

## Abstract

**Background:** Yellow fever remains a public health threat in Cameroon, especially in conflict-affected regions where surveillance systems face contextual challenges. Community Health Workers (CHWs) are crucial in early detection and response under the Integrated Disease Surveillance and Response (IDSR) framework. Understanding their perceptions is essential to improve performance.

**Methods:** This qualitative study explored CHWs’ and health personnel’s perspectives on yellow fever surveillance in two districts of Northwest Cameroon. Data were collected through seven focus group discussions and fifteen in-depth interviews. Thematic analysis followed Braun and Clarke’s framework using MAXQDA 2022.

**Results:** Six themes emerged: (1) mixed understanding of yellow fever among communities; (2) trust in CHWs shaped by social embeddedness; (3) emotional and professional burdens; (4) limited feedback and supervision; (5) cultural and spiritual barriers; and (6) gender-specific experiences. CHWs were central to surveillance yet often under-recognized.

**Conclusions:** CHWs are vital but under-supported in yellow fever surveillance. Although trusted by communities, their work is hindered by limited training, weak feedback systems, and sociocultural dynamics. Strengthening support, integrating gender-sensitive strategies, and engaging with local beliefs are necessary to improve IDSR outcomes in fragile health settings.

## Introduction

Community Health Workers (CHWs) have been globally recognized for their pivotal role in delivering essential health services, particularly in low- and middle-income countries (LMICs) where healthcare access remains limited [1]. As trusted members of the communities they serve, CHWs bridge the gap between formal healthcare systems and underserved populations by providing health education, promoting disease prevention, and facilitating access to primary care services [2,3]. In regions heavily burdened by communicable diseases, CHWs are often the first point of contact for early disease detection, outbreak reporting, and referral [4]. The World Health Organization (WHO) introduced the Integrated Disease Surveillance and Response (IDSR) strategy to strengthen public health surveillance systems across the African region [5]. IDSR aims to integrate multiple disease surveillance activities into a unified, efficient, and effective approach that emphasizes prompt detection, verification, reporting, and response to priority diseases. Within this framework, CHWs play an indispensable role in promoting community participation, collecting surveillance data, sensitizing communities on disease prevention, and serving as the first responders to health threats [6,7]. Evidence indicates that robust CHW programs significantly improve health indicators such as vaccination coverage, maternal and child health outcomes, and reduction in preventable diseases [8,9]. However, despite their critical role, CHWs face persistent barriers that undermine their performance and the overall effectiveness of surveillance systems. Notably, limited access to essential resources, including surveillance registers, transportation, and communication devices, continues to constrain CHWs’ ability to perform their duties efficiently [10,11]. Inadequate or irregular training further exacerbates these challenges by leaving CHWs ill-equipped to identify, document, and report disease outbreaks according to standardized protocols [12]. Community perceptions and misconceptions about disease transmission also pose significant challenges to CHWs’ efforts in engaging communities effectively [13]. In many settings, traditional beliefs and misinformation about the causes of infectious diseases create resistance to public health interventions, complicating efforts to promote surveillance and preventive practices [14]. Addressing these misconceptions through culturally appropriate health education and continuous community engagement is essential for improving public trust and cooperation in disease surveillance efforts [24]. Fragmentation between community-based surveillance and formal healthcare systems further limits the responsiveness and efficiency of public health surveillance. Weak coordination, inconsistent reporting pathways, and limited integration of CHWs into facility-based referral and feedback loops contribute to delays in case detection and response. Building stronger linkages between CHWs, healthcare workers, and district surveillance teams is vital for improving timely disease reporting and enhancing system-wide responsiveness [18,22]. Given the limited integration of CHWs into formal surveillance systems and the complex context of the Northwest Region of Cameroon, there is a need to better understand the challenges that may be affecting their engagement with communities in disease surveillance activities. This is particularly critical in areas like the Bali and Bamenda Health Districts, where insecurity, resource constraints, and social disruption may further weaken surveillance functions [19,20,23]. This study seeks to explore the barriers that CHWs and frontline health personnel face in carrying out IDSR-related responsibilities, based on their perspectives. While community perceptions are not directly captured, this study examines how CHWs and health workers interpret and respond to community behaviors related to yellow fever, thereby informing strategies to strengthen community-level surveillance and public health response.

## Materials and methods

### 2.1 Study design and setting

This study employed an exploratory qualitative research design to investigate CHW and health personnel perceptions regarding yellow fever surveillance and response under the IDSR framework. The study was conducted in the Bamenda 3 and Bali health districts of Cameroon’s Northwest Region areas characterized by ongoing socio-political unrest, internal displacement, and weakened health infrastructure. These contextual dynamics significantly influence community trust in formal health interventions and the functionality of disease surveillance systems. Ethical approval for this study was obtained from the Institutional Review Board of the University of Bamenda (Ref: 2024/0006H/UBa/IRB), approved on 12 January 2024. All procedures were performed in accordance with the Declaration of Helsinki.

### 2.2 Participants and Sampling

Participants were purposefully selected from two health districts (Bamenda 3 and Bali) based on their active roles in yellow fever surveillance. The study involved 24 CHWs who participated in seven focus group discussions (FGDs), and 15 frontline health workers and surveillance officers who participated in in-depth interviews (IDIs). No CHW describing community beliefs were recruited directly. All participants were engaged in routine surveillance activities and were familiar with the IDSR system. This sampling allowed for triangulation across different cadres of health actors engaged in surveillance.

Participants were recruited between 15 January 2024 and 30 March 2024.

### 2.3 Data Collection

Data were collected using semi-structured guides tailored for FGDs with CHWs and IDIs with health staff. FGDs allowed CHWs to discuss shared challenges, community interactions, and surveillance practices, while IDIs enabled deeper insight into the institutional and operational challenges faced by health personnel. Interviews were conducted in English or Pidgin English and were audio recorded with participant consent. Field notes were also maintained. All data were transcribed verbatim, and identifiers were removed to ensure confidentiality. Written informed consent was obtained from all participants prior to participation. For participants younger than 18 years, parental or guardian consent was obtained. The consent process was reviewed and approved by the University of Bamenda Institutional Review Board.

### 2.4 Data management and analysis

Recordings were transcribed verbatim and translated where necessary. Transcripts were de-identified, coded using MAXQDA 2022, and analyzed thematically following Braun and Clarke’s six-step framework. Coding was iterative and inductive, beginning with open coding and evolving into focused coding through constant comparison. Thematic saturation was monitored across transcripts, and data were triangulated between IDIs and FGDs. Two researchers independently coded a subset of transcripts and discussed discrepancies until consensus was reached, enhancing reliability and consistency.

All transcripts were anonymized, and no personally identifiable information was collected. Data were stored securely and were accessible only to the research team.

## Results

A total of 135 participants were included in the study, comprising 120 Community Health Workers (CHWs) who participated in 12 Focus Group Discussions (FGDs) one per health area across the two districts (7 in Bali and 5 in Bamenda 3). Each FGD consisted of 10 purposively selected CHWs involved in IDSR and community vaccination activities. In addition, 15 in-depth interviews (IDIs) were conducted with CHWs and local health stakeholders, selected to ensure variation in roles, gender, years of experience, and geographic distribution across urban and rural communities within the Bamenda 3 and Bali health districts. Thematic analysis generated six core themes that capture participants’ perceptions and lived experiences in yellow fever surveillance under the IDSR framework.

### Understanding of yellow fever and disease perception

Participants reported mixed understanding of yellow fever within the communities. While a few recognized it as a mosquito-borne viral disease, most associated it with witchcraft, punishment from ancestors, or spiritual attack due to unfamiliar symptoms like jaundice and sudden death. CHWs noted that these misconceptions often led to late reporting, fear, and reliance on traditional healers rather than health facilities. “When they see yellow eyes or someone suddenly weak and vomiting, they say it is not a hospital sickness… they say someone has been bewitched,” said a CHW from Bali. This distorted perception made it difficult for CHWs to mobilize timely response or convince families to accept referral and vaccination.

### Community Trust and Attitudes Towards CHWs

Trust in CHWs was generally high, especially when the CHW was a long-time resident and known to the community. Familiarity allowed CHWs to navigate sensitive situations and encourage surveillance cooperation. However, in areas where CHWs were newly posted or perceived as affiliated with political actors, community suspicion hindered their work. “Some think we are agents of the government, and in this crisis, that can be dangerous,” noted one CHW. Trust was also found to be relational, built over time through presence, responsiveness, and respectful engagement.

### Perceived role and professional identity of CHWs

CHWs identified strongly with their role as the first line of public health surveillance. They viewed themselves as “the eyes of the community” and felt pride in contributing to outbreak detection. However, many expressed frustrations at the lack of institutional recognition, describing themselves as “invisible” despite their workload. “We are the ones walking from house to house, yet our names are not on any list when trainings or allowances come,” remarked a CHW from Bamenda. This perceived marginalization negatively impacted morale and retention.

### Surveillance Reporting Experience and Feedback Gaps

While CHWs understood the procedures for reporting suspected cases, they highlighted inconsistent feedback and supervision from the health system. Several described submitting reports weekly without acknowledgment or any follow-up support. “You keep reporting and nothing comes back… it feels like no one is listening,” a CHW commented. The absence of responsive feedback loops discouraged proactive case detection and undermined the value of surveillance work.

### Cultural and emotional dimensions of surveillance work

CHWs frequently navigated complex cultural landscapes when conducting surveillance. They had to be tactful in discussing illness, particularly in families with strong traditional or spiritual beliefs. Some CHWs reported emotional stress when accused them of bringing misfortune or spying for authorities. “I cried one day when a woman insulted me and said I wanted to bring death to her family,” recounted a female CHW. Despite this, many CHWs used coping strategies such as peer support, prayer, and reflection to manage the emotional burden.

### Gender-specific experiences

Gender roles significantly influenced how CHWs experienced their surveillance tasks. Female CHWs were more accepted into households for maternal and child health discussions but were sometimes restricted from entering certain male-dominated settings or traveling alone. “As a woman, I can enter most homes… but I cannot go alone to some quarters when it is late,” shared a CHW. In contrast, male CHWs often took on leadership in outbreak response but faced expectations of physical mobility and assertiveness. These gendered experiences shaped engagement styles and risks faced in the field.

## Discussion

This qualitative study explored how and CHWs perceive yellow fever surveillance and how those perceptions influence engagement with the IDSR system. Findings underscore that disease surveillance is not simply a technical or data-driven process but one deeply shaped by social, cultural, emotional, and institutional dynamics. These insights are particularly critical in fragile health systems affected by conflict and mistrust.

### Disease perception and cultural interpretations

A key finding was the widespread community misperception of yellow fever as a spiritual or supernatural illness. These beliefs significantly hinder early case detection and healthcare-seeking. Such perceptions have been reported in other African contexts with ambiguous or sudden illnesses like Ebola or polio, where traditional interpretations often take precedence over biomedical understanding [24,25]. In Sierra Leone during the Ebola epidemic, misinterpretation of symptoms led to widespread resistance to surveillance and treatment [24]. Similarly, in northern Nigeria, yellow fever and polio were often interpreted as government conspiracies [25]. These studies highlight the need for surveillance communication strategies to engage local health worldviews directly.

### Trust and the relational role of CHWs

CHWs were both respected and mistrusted depending on their community integration. This duality is consistent with findings by Perry et al., who emphasized that CHW effectiveness depends on their embeddedness within local social networks [26]. Trust in fragile contexts is built relationally over time. Where it is absent, particularly during political crises, surveillance is weakened. Qualitative studies in Uganda and Liberia confirm that when CHWs are not perceived as ‘belonging’ to the community, data quality and community participation decline [27,28].

### Surveillance works as emotional and invisible labor

Surveillance Work as Emotional and Invisible Labor CHWs described surveillance as emotionally draining and socially unrecognized what has been called “invisible labor” in global health literature [29]. In Ethiopia and Mozambique, community health volunteers similarly described their contributions as undervalued despite their central role in community-level services [30]. The emotional strain, lack of recognition, and absence of mental health support are well-documented reasons for attrition in community health programs [31]. This reinforces the need for psychological and social support mechanisms alongside technical training.

### Reporting gaps and feedback loops

Reporting Gaps and Feedback Loops The lack of feedback described by CHWs is a well-known weakness in decentralized surveillance systems. Studies in Ghana and Tanzania reported similar frustrations among CHWs, where one-way reporting led to demotivation and reduced vigilance [32,33]. Bi-directional communication via feedback calls, peer meetings, or visual dashboards has been shown to improve surveillance motivation [34].

### Gendered dynamics in surveillance engagement

Gendered Dynamics in Surveillance Engagement Gender significantly shaped CHWs’ access, trust, and safety. Women were often better positioned to engage households, particularly in maternal and child contexts, but faced mobility and safety constraints. Similar patterns have been documented in India and Kenya, where female CHWs were restricted from conducting night visits or entering male-dominated spaces [35,36]. Programs that fail to integrate gender-sensitive strategies in CHW design risk undermining participation and effectiveness [37].

### Policy and programmatic implications

Policy and Programmatic Implications Our findings call for a rethinking of how surveillance systems are built and sustained. CHWs need training in communication, negotiation, and trauma-informed care, not just reporting protocols. Feedback mechanisms and visible recognition must be incorporated into routine surveillance workflows [38]. Moreover, gender and cultural competence should be prioritized in recruitment, deployment, and supervision policies to ensure equitable and effective surveillance reach [39].

### Study limitations

While the study provides deep insight into perceptions and lived experience, its qualitative design limits generalizability. Conflict dynamics may have influenced responses, and some depth may have been lost in translation from Pidgin English. However, triangulation across FGDs and IDIs, and thematic saturation, support the credibility of the finding

## Conclusion and recommendations

This study reveals that CHWs and health personnel perspectives and CHW experiences profoundly shape the success of yellow fever surveillance under the IDSR strategy in fragile settings like Northwest Cameroon. Rather than being passive reporters, CHWs act as trusted intermediaries, cultural translators, and emotional laborers operating at the frontline of epidemic detection. Their effectiveness is deeply dependent on the social, cultural, and emotional environment in which they operate. Key findings demonstrate that disease perceptions in the community are not always aligned with biomedical understanding, often leading to delayed case reporting and vaccine hesitancy. CHWs, while largely trusted, face relational challenges when operating in new or politically sensitive communities. They report feeling invisible within formal systems despite their contributions, and the absence of responsive feedback mechanisms undermines morale and commitment. Gender dynamics further influence CHW engagement, with women navigating restricted mobility but benefiting from trust in caregiving roles. To strengthen surveillance effectiveness in such contexts, we recommend the following:

Enhance CHW Cultural Competence: Training should go beyond clinical tasks to include modules on local health beliefs, communication strategies, and conflict-sensitive engagement. Institutionalize

Feedback Mechanisms: Surveillance systems must implement simple, regular two-way communication channels, such as SMS feedback, community review meetings, or supervisory visits to validate reports and boost morale.

Recognize CHW Contributions: Establish visible and symbolic recognition systems—certificates, awards, or radio mentions alongside material incentives to affirm the value of CHWs’ work.

Apply Gender-Responsive Surveillance Design: Deploy CHWs in a manner that aligns with cultural norms while ensuring that gender-related risks and barriers are proactively addressed through supervision and peer support.

Integrate Community Voices in Surveillance Planning: Engage community leaders and traditional healers in surveillance sensitization to bridge the biomedical-local knowledge divide and foster community trust in IDSR systems.

Provide Emotional Support for CHWs: Regular debriefing sessions, peer support networks, and access to psychosocial services can help manage the emotional toll of frontline surveillance work.

## Data Availability

Data cannot be shared publicly because they contain information that could compromise participant privacy. The data are available from the Institutional Review Board of the University of Bamenda (Ref: 2024/0006H/UBa/IRB) for researchers who meet the criteria for access to confidential data

---

**Table 1:** Sociodemographic Characteristics of Study Participants (N=135)

| Variable | CHWs (n=120) | Health Staff (n=15) |
| --- | --- | --- |
| Age Range (years) | 25–60 | 30–58 |
| Gender | 68 Female, 52 Male | 6 Female, 9 Male |
| Education Level | Secondary–Tertiary | Tertiary |
| Years of Experience | 2–15 years | 5–20 years |
| Health District | Bamenda 3 & Bali | Bamenda 3 & Bali |

**Table 2:** Thematic Summary with Illustrative Quotes.

| Theme | Illustrative Quote |
| --- | --- |
| Mixed Understanding of Yellow Fever | ‘Some people think it is just malaria, others believe it is a punishment from the ancestors’ (FGD, CHW). |
| Trust in CHWs | ‘We listen to them because they are our people, and they know our problems’ (IDI, health staff). |
| Invisibility of CHW Work | ‘We do all the work, but nobody even says thank you’ (FGD, CHW). |
| Feedback Gaps | ‘We report cases but never hear what happens next’ (IDI, CHW). |
| Cultural and Spiritual Barriers | ‘Some believe yellow fever is caused by witchcraft and don’t come to the hospital’ (FGD, community member). |
| Gender-Specific Challenges | ‘As a woman, I cannot go out at night to do follow-up, it is not safe’ (FGD, female CHW). |

